# Circulating microglia-derived extracellular vesicles predict recovery after rehabilitation in stroke survivors

**DOI:** 10.1101/2025.11.25.25341029

**Authors:** Aurora Mangolini, Silvia Picciolini, Pietro Arcuri, Donata Bardi, Leandro Cosco, Francesca Cecchi, Lorenzo Romagnoli, Ester Marra, Pietro Parisse, Andrea Mannini, Jorge S. Navarro, Marzia Bedoni, Alice Gualerzi

## Abstract

**Background:** Timely intensive rehabilitation is crucial to contrast the negative escalation of events that follow a stroke injury, to promote tissue regeneration, and to restore the physiological function. This study aimed to identify measurable blood biomarkers that could be predictive of functional recovery in stroke survivors.

**Methods:** 30 sub-acute stroke subjects were enrolled at Fondazione Don Carlo Gnocchi Onlus (Italy) during the observation prospective cohort study EXO4STROKE (NCT05370105). Among the clinical scales used for patient profiling, modified Barthel Index (MBI) was considered as primary outcome for the evaluation of rehabilitation recovery. Cytokines, Neurofilaments and circulating Extracellular Vesicles (EVs) were evaluated in the serum of patients at admission (T0) and at discharge (T1) in intensive rehabilitation unit. The Surface Plasmon Resonance imaging (SPRi) technique was exploited to obtain a multiplexed analysis of circulating EVs from brain (neurons, astrocytes, microglia) and non-brain (endothelium, skeletal muscle and platelets) cells, and for the relative quantification of markers of pathological or regenerative processes. Machine learning-based analysis complemented the study by integrating statistically significant features in a cross-validated prediction model targeting functional recovery at T1 from T0 data.

**Results:** High serum levels of IL-6 and Neurofilament Light Chain at T0 were associated with higher residual disability (lower outcome) at T1. In contrast, elevated circulating levels of EVs from neurons (CD171+) and microglia (CD11b+) were associated with better recovery after rehabilitaion. Moreover, the overexpression of the VEGF receptor on microglial EVs (IB4+ and CD11b+), was associated with higher functional improvement. Alternatively, the overexpression of TGF-β receptor on IB4+ EVs during the subacute phase post-stroke was found associated with increased stroke severity. The MBI at T1 was predicted with a median [iqr] absolute error of 5.7[8.7] points.

**Conclusion:** This study identifies a panel of few circulating biomarkers associated to Evs able to objectively assess post-stroke status and predict functional recovery after rehabilitation.

## Introduction

Stroke is the leading cause of death and disability worldwide: 1 in 4 adults over the age of 25 will experience a stroke. Over 12 million people worldwide have a first stroke each year and 7 million, while most survivors face devastating cognitive, motor, and functional impairments ^1,2^. After the acute injury (ischemic or haemorrhagic) and neuronal cell loss caused by alteration of blood circulation, a secondary neurodegenerative process may evolve days, weeks, and even months after the event^3^. Recovery after stroke is generally classified into i) neurological recovery, dependent on lesion type and location, and ii) functional recovery, influenced by environment, rehabilitation, and patient motivation ^4^. Timely and intensive rehabilitation is crucial to counteract the cascade of detrimental events that follow the injury and promote a positive niche for the regeneration, tissue repair and functional restoration^5^.

Clinical scales are currently used to guide rehabilitation. Despite being useful tools for guiding decisions in acute and chronic management, clinical scales might oversimplify patients’ conditions and fail to fully capture the impact of neurological deficits. In this scenario, inflammation shows a detrimental time-dependent role, perpetuating the injury, but also contributing to tissue regeneration ^6^. Moreover, the different components of the neurovascular unit, i.e. neurons, microglia, astrocytes and vascular cells, show distinct responses to ischemia and inflammation.

Biomarkers measurable in accessible biofluids could therefore offer a more objective means to characterize patients, clarify the mechanisms activated after stroke, and support personalized rehabilitation strategies.

To date, many candidates have been investigated, including inflammatory mediators, tissue-derived cytokines, growth factors, hormones, neurofilaments, and microRNAs, but none have yet reached clinical practice and most of them are still under investigation ^7^. Biomarker levels are strongly influenced by the integrity of the blood-brain barrier and, a single analyte may not adequately represent the heterogeneous and dynamic nature of stroke.

In recent years, Extracellular Vesicles (EVs) have emerged as promising biomarkers. EVs are lipid bilayer nanostructures released from all cells, carrying biologically active molecules (including proteins, lipids, and extracellular RNAs) and capable of eliciting recipient-cell response ^8^. EV biochemical composition reflects the physiological or pathological state of the parent cell and their ability to cross most anatomical barriers, including the blood-brain barrier, makes them valuable candidates for probing brain-related processes ^5^.

Several neural and circulating cell types release EVs into the blood after stroke ^9^. Although their diagnostic potential has been widely explored, their prognostic value and their role in functional recovery remain less investigated.

The present work aims to examine how rehabilitation influences different EV populations and to evaluate their surface molecules as potential predictive biomarkers of post-stroke recovery. To reach these purposes, a multiplexed, highly sensitive biosensor was used to detect and characterize multiple EV populations. In parallel, the inflammatory profiling of patients was performed by measuring circulating cytokines and soluble factors involved in neuroinflammation and neurodegeneration.

## Methods

### Patient Recruitment and Samples Collection

The EXO4STROKE trial (ClinicalTrials ID: NCT05370105) was a prospective case-control trial approved by Fondazione Don Carlo Gnocchi Ethical Committee (protocol number: 20/2018/CE_FdG/FC/SA) in 2018 and completed in July 2023. The present work reports the longitudinal observation of 30 ischemic or hemorrhagic stroke patients recruited at IRCCS Santa Maria Nascente (Milan, IT) and IRCCS Don Gnocchi (Florence, IT). All participants or their representatives provided written informed consent according to the Declaration of Helsinki. Patients entered the rehabilitation program 15-30 days after stroke (subacute phase). Exclusion criteria included age < 35 years and > 75 years, relapse, traumatic brain injury, concomitant or past diagnosis of oncological, immunological, haematological and neurodegenerative diseases.

Clinical assessment included Modified Barthel Index (MBI), Modified Rankin Scale (MRS), Cumulative Illness Rating Scale (CIRS), Numerical Rating Scale (NRS), National Institutes of Health Stroke Scale (NIHSS), Mini-Mental State Examination (MMSE), Fugl-Meyer Assessment (FMA) scale, Trunk control test, Short Physical Performance Battery (SPPB) test, Functional ambulation classification (FAC) test, Hospital Anxiety and Depression Scale (HADS), and Modified Ashworth Scale (MAS). Patients were evaluated at admission (T0) and at discharge (T1), i.e., after completing the assigned rehabilitation protocol. All data were collected and managed using REDCap electronic data capture tools ^10^.

Integrated Rehabilitation Pathway was based on the AHS/ASA guidelines ^11,12^ and national requirements, providing at least 3 hours/day of specific rehabilitation interventions (physiotherapy, cognitive rehabilitation, speech and dysphagia therapy, occupational therapy, training in the use of aids, psychological support). Physiotherapy may also include robotic rehabilitation according to the individual rehabilitation plan defined by the interdisciplinary team. The rehabilitation plans were based on the assessment at admission and adapted to emerging needs during the rehabilitation stay, through systematic weekly team meetings and on demand.

Blood samples (10 ml) were collected at T0 and T1. Serum was obtained by centrifugation (10 minutes, 2500 x *g* at room temperature), aliquoted in cryovials, anonymized and stored at –80 °C until use. Haemolysis risk was assessed by absorbance at 414 nm (Nanodrop, UV-VIS analysis)^13^, as suggested by the international guidelines for blood-derived EV isolation ^14^ (Supplementary Material).

### Enzyme-Linked Immunosorbent Assay on Serum Samples

Inflammatory cytokines (Tumor Necrosis Factor alpha (TNF-α), Interleukins 6 and 10 (IL-6, IL-10), Fas Ligand (FasL)), Brain Derived Neurotrophic Factor (BDNF), Vascular Endothelial Growth Factor Receptor 2 (VEGFR2), and peptide hormone Leptin were quantified in 28 subjects before (T0) and after (T1) rehabilitation, while Neurofilament Light chain (NFL) and Neurofilament High chain (NFH) were quantified in 30 patients at T0 and T1.

### Extracellular Vesicles

Details about the EV isolation, EV characterization, and SPRi biochip preparation are reported in Supplementary Material.

EVs were isolated from 500 µL of serum by size exclusion chromatography. To evaluate the size distribution and the concentration of the isolated EVs, Nanoparticle Tracking Analysis (NTA) was performed. Reported markers of small EVs (CD9, TSG 101, ALIX, Flotillin-1), and EVs from neurons (CD171), endothelium (VCAM-1), skeletal muscle (Irisin), microglia (CD11b), astrocytes (GLAST), and platelets (CD41) were evaluated by Dot blot assay. Atomic Force Microscopy (AFM) analysis was performed to evaluate the physical and biomechanical properties of EV samples following protocol proposed by Ridolfi et al. ^15^

An optimized SPRi biosensor was used to characterize the populations of serum EVs in people before and after the rehabilitation therapy. The biosensor was functionalized to recognize simultaneously EVs subfamilies coming from skeletal muscle cells (Irisin+), endothelium (CD106+, CD31+), platelets (CD41+), microglia (CD11b+, IB4 lectin+), neurons (CD171+), and astrocytes (GLAST+). Additional ligands spotted on the biosensor surface were Ab anti-Human Klotho as a marker of aging, Ab anti-Human CD9 as a general marker of EVs, and anti-Human Transforming Growth Factor beta Receptor II (TGF-βRII) to evaluate the involvement of vesicles in inflammation signalling. Secondary labelling was also performed by injecting PK-11195, TGF-β1, and VEGF with the aim of assessing the expression of the corresponding receptors (TSPO, TGF-βR and VEGFR) on the membrane of the immobilized EVs.

### Statistical Analysis

Descriptive statistical analysis was performed using OriginPro2023b. For numerical variables, the mean, median, standard deviation, and interquartile range were calculated, while for categorical variables, relative frequencies were computed. The Wilcoxon signed-rank non-parametric test was used to evaluate the statistical difference between T0 and T1 data obtained from ELLA, NTA, BCA assay and SPRi data. To evaluate the possible correlations between clinical scales and the inflammatory soluble factors and the proposed EV-associated biomarker, a partial correlation study was performed, using age, sex and time between acute event and admission to rehabilitation program as controlling variables. In this regard, the variation of the clinical parameters was considered defining the balanced variation in the clinical scores with the following formula: ΔSCALE= [(SCALEscore *at* T1 – SCALEscore *at* T0)/ SCALEscore *at* T0)]. Since the descriptive analysis revealed a non-normal distribution of the data, Spearman’s tests were applied for correlation analyses. The significance level α was set at 0.05 for all tests. Machine learning-based analysis was then used to examine the association between the MBI total score at T1 and concentration-related variables, including inflammatory, endothelial, and neuronal biomarkers. Details are reported in Supplementary Material.

## Results

The major demographic and clinical features of the selected cohort are summarized in Table 1. MBI score was considered as primary outcome for the evaluation of rehabilitation recovery.

**Table 1:** Main demographic and clinical parameters of stroke subjects at admission (T0) and discharge (T1). All data are reported as mean value ± 1 standard deviation (SD).

| *n=30* | *mean ± 1SD* |  |
| --- | --- | --- |
| Age (years) | 66.47 ± 9.88 | |
| Gender | 19 male/ 11 female |  |
| Time between stroke event and enrolment (days) | 20.43 ± 12.53 | |
| Rehabilitation period (days) | 41.3 ± 15.63 | |
| Stroke Type | 25 ischemic/5 hemorrhagic |  |
|  | **T0** | **T1** |
| MBI | 44.9 ± 28.76 | 80.59 ± 18.88 |
| MMSE | 26.24 ± 2.99 | 26.16 ± 4.74 |
| NIHSS | 5.07 ± 3.43 | 2.77 ± 2.65 |
| CIRS | 20.17 ± 4.72 | 20.17 ± 4.72 |
| NRS | 1.32 ± 2.07 | 1.17 ± 2.90 |
| MRS | 3.87 ± 0.69 | 2.5 ± 1.14 |

### Biomolecular profile

Selected inflammatory cytokines and soluble factors currently assigned a role in stroke pathophysiology and patient profiling^7^ were quantified using the ELLA platform (Table 2).

**Table 2:**
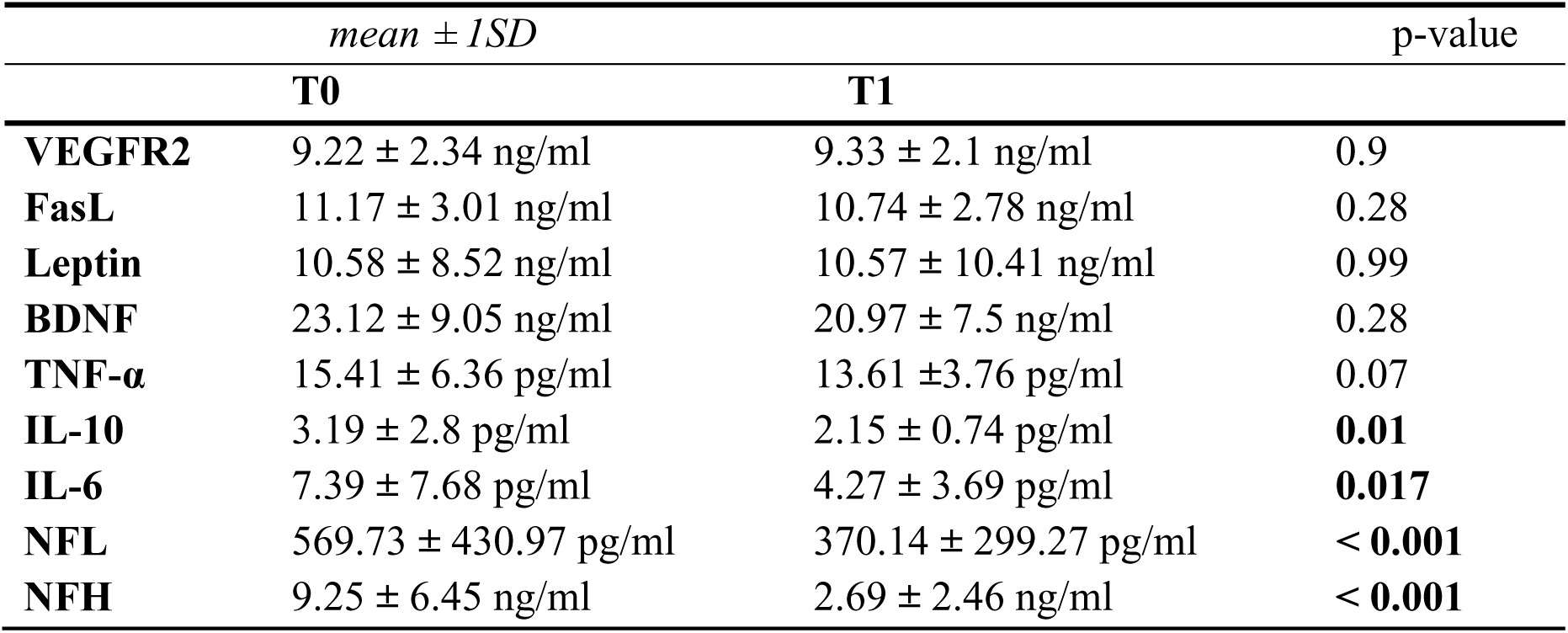
Summary of serum concentration of inflammatory cytokines and soluble factors measured by enzyme-linked immunosorbent assay before (T0) and after (T1) the rehabilitation program. All data are reported as mean value ± 1 standard deviation (SD). The Wilcoxon signed-rank non-parametric test was used to compare the distribution of the two time points.

|  | *mean ± 1SD* |  | p-value |
| --- | --- | --- | --- |
|  | **T0** | **T1** |  |
| **VEGFR2** | 9.22 ± 2.34 ng/ml | 9.33 ± 2.1 ng/ml | 0.9 |
| **FasL** | 11.17 ± 3.01 ng/ml | 10.74 ± 2.78 ng/ml | 0.28 |
| **Leptin** | 10.58 ± 8.52 ng/ml | 10.57 ± 10.41 ng/ml | 0.99 |
| **BDNF** | 23.12 ± 9.05 ng/ml | 20.97 ± 7.5 ng/ml | 0.28 |
| **TNF-α** | 15.41 ± 6.36 pg/ml | 13.61 ± 3.76 pg/ml | 0.07 |
| **IL-10** | 3.19 ± 2.8 pg/ml | 2.15 ± 0.74 pg/ml | **0.01** |
| **IL-6** | 7.39 ± 7.68 pg/ml | 4.27 ± 3.69 pg/ml | **0.017** |
| **NFL** | 569.73 ± 430.97 pg/ml | 370.14 ± 299.27 pg/ml | **< 0.001** |
| **NFH** | 9.25 ± 6.45 ng/ml | 2.69 ± 2.46 ng/ml | **< 0.001** |

No significant differences were observed for VEGFR2, FasL and Leptin (Figure 1A, 1B, 1C) comparing levels before and after rehabilitation. Similarly, BDNF and TNF-α levels were slightly decreased after rehabilitation, despite no statistically significant variation (Figure 1D, 1E). In contrast, **significant decrease of the expression of IL-10 and IL-6** were observed at discherge (Figure 1F, 1G).

**Figure 1:**
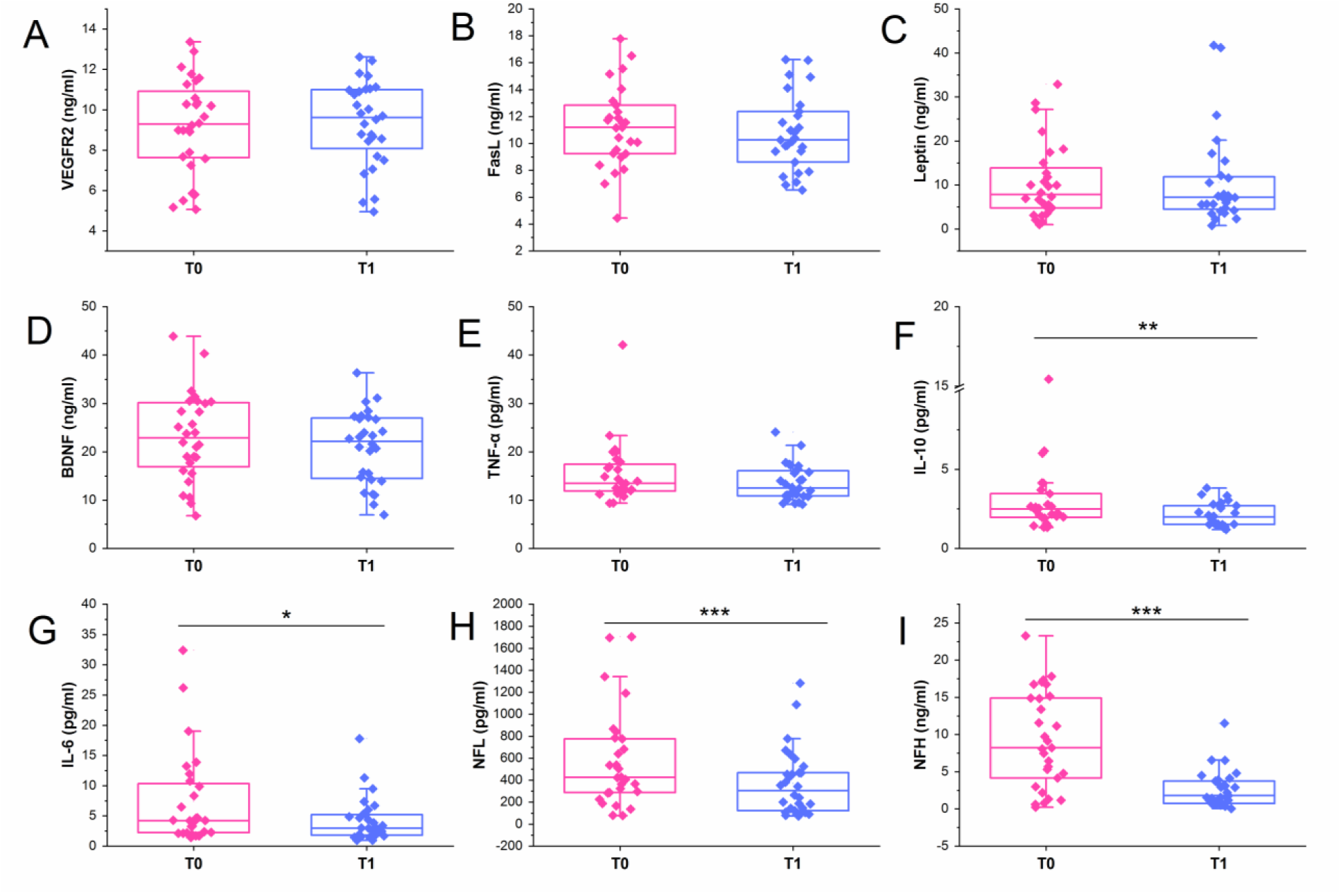
Stroke patients biomolecular profiling. Box plots represent the quantification of inflammatory cytokines and soluble factors by enzyme-linked immunosorbent assay on serum samples of stroke patients (*n=*28 for VEGFR2, FasL, Leptin, BDNF, TNF-α, IL-10, IL-6; *n=*30 for NFL and NFH) before (T0) and after (T1) the rehabilitation program. The results obtained for VEGFR2 (**A**), FasL (**B**), Leptin (**C**), BDNF (**D**), TNF-α (**E**), IL-10 (**F**), IL-6 (**G**), NFL (**H**), NFH (**I**) are reported. The Wilcoxon signed-rank non-parametric test was used to compare the distribution of the two time points.*p<0.05; **p<0.01; ***p<0.001.

Neurofilaments levels showed **a remarkable decrease of NFL** and NFH serum concentration after rehabilitation (Figure 1H and 1I), although remaining above the typical control range of the age and sex matched population (data not shown).

### EV Physico-Chemical Characterization

NTA demonstrated slightly higher EV concentration, size, and in the amount of protein per particle at T1 compared to T0 although differences were not statistically significant after paired sample Wilcoxon signed rank test (Table 3, Figure S1). Dot Blot assay confirmed the EV nature of the isolated particles by assessing the presence of markers for small-EV markers (CD9, TSG 101, ALIX, Flotillin-1), neurons (CD171+), endothelium (VCAM-1+), muscle (Irisin+), microglia (CD11b+), astrocytes (GLAST+), and platelets (CD41+) (Figure S2).

**Table 3:**
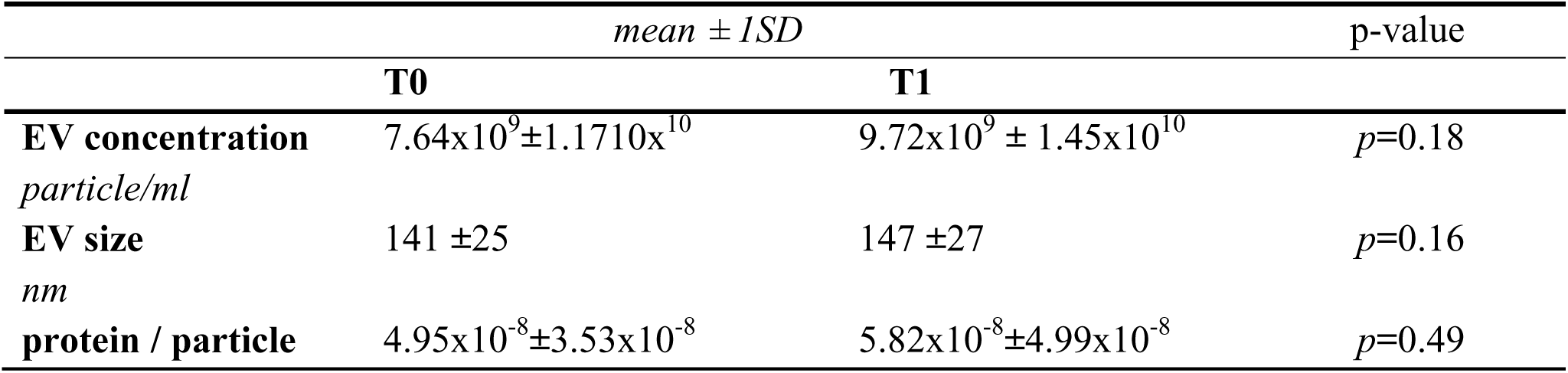
Summary of EV concentration, size and protein per particle amount obtained by Nanoparticle Tracking Analysis (NTA) and BCA assay. p values are referred to non parametric paired sample Wilcoxon signed rank test.

AFM imaging showed comparable morphology and biomechanics between T0 and T1, with a greater number of vesicles observed at T1. In Figure 2, AFM topographic images represent T0 and T1 samples. In the 5µm x 5µm AFM scans (Figure 2A, 2D) we clearly observed the presence of roundish particles whose size was compatible with the expected dimensions of EVs (30-200 nm), as evidenced by the high-resolution images (Figure 2B, 2E) and corresponding height profiles (Figure 2C, 2F). Looking at size distribution, AFM confirmed the NTA observation: the two samples are very similar, as evidenced in the height vs equivalent diameter scatter plot and in the box plot reported in Figure 2 with people at T1 presenting an increased number of vesicles attached to the surface (5-6 per scan for T0, 15 per scan for T1). The contact angle vs equivalent diameter dispersion reported in Figure 2H also show no significant difference in the overall behaviour of EVs, indicating similar biomechanical properties.

**Figure 2:**
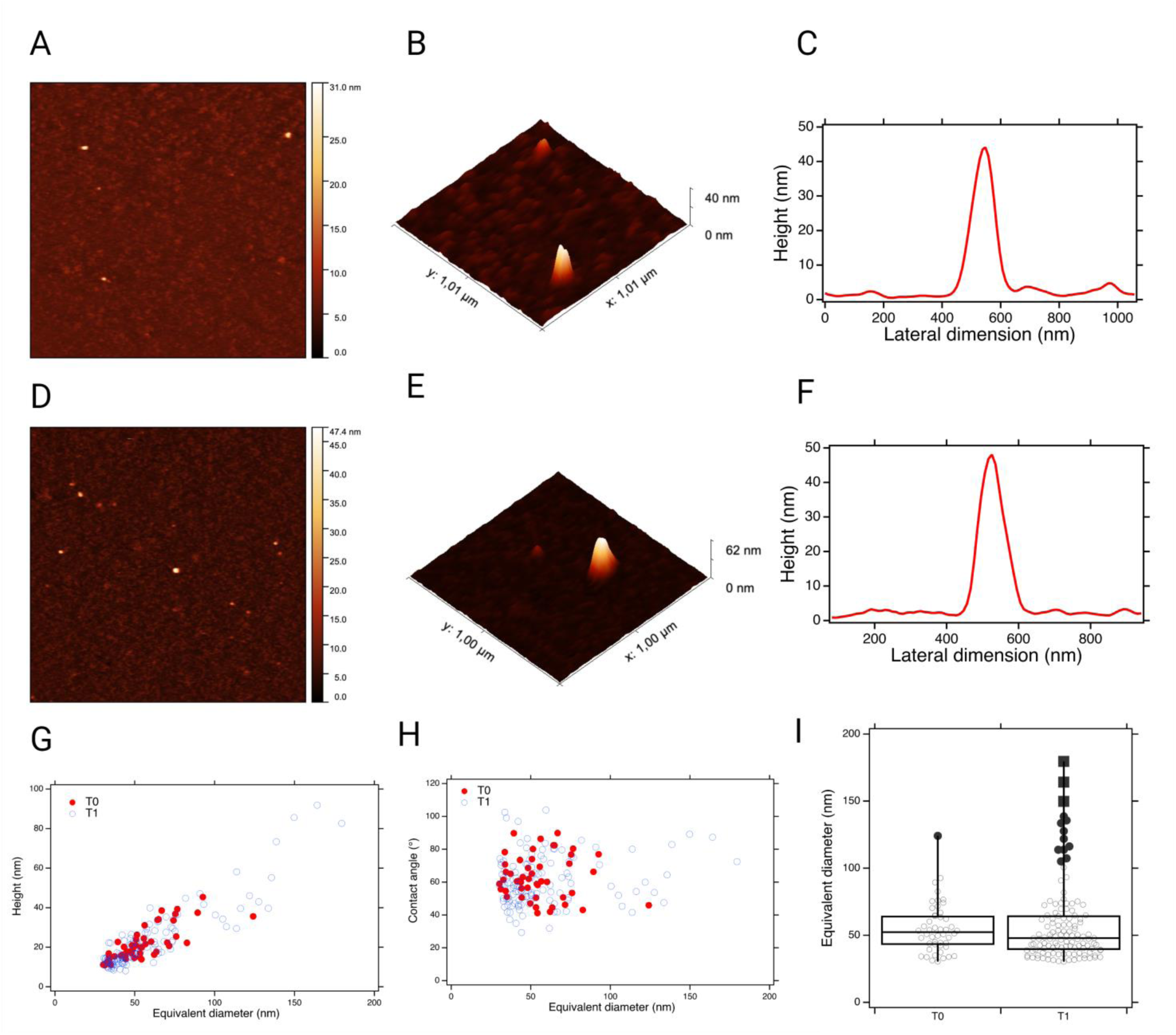
5µm x 5µm AFM topographic scan, 1mm x 1mm 3d rendering AFM topographic scan and corresponding line profiles respectively for T0 (**A,B,C**) and T1 (**D,E,F**). Height vs equivalent diameter scatter plot for T0 (red dots) and T1 (Blue circles) samples (**G**). Contact angle vs equivalent diameter scatter plot for T0 (red dots) and T1 (Blue circles) samples (**H**). Box plot of the equivalent diameter distributions for T0 and T1 samples (**I**).

### SPRi analysis

SPRi signal intensities collected at the end of the injection of EVs were indicative of the relative amount of each EV population. The detection of all the subfamilies (both brain and non-brain derived EVs) was successful in all samples despite considerable inter-individual variability (Figure S3B). Overall, the relative amount of EV subfamilies showed variability between T0 and T1, without a statistical significance. Considering the differences of SPRi signals collected on ligand spots (Δ = T1 SPRi intensity – T0 SPRi intensity), we observed a reduction of EV populations coming from astrocytes (GLAST+), skeletal muscle cells (Irisin+), and Klotho+ EVs at T1. On the other hand, the rehabilitation treatment seems to increment the amount of circulating EVs from microglia (IB4+, CD11b+), platelets (CD41+), neurons (CD171+), endothelium (CD106+), and TGF-βRII+ EVs (Figure 3A).

**Figure 3:**
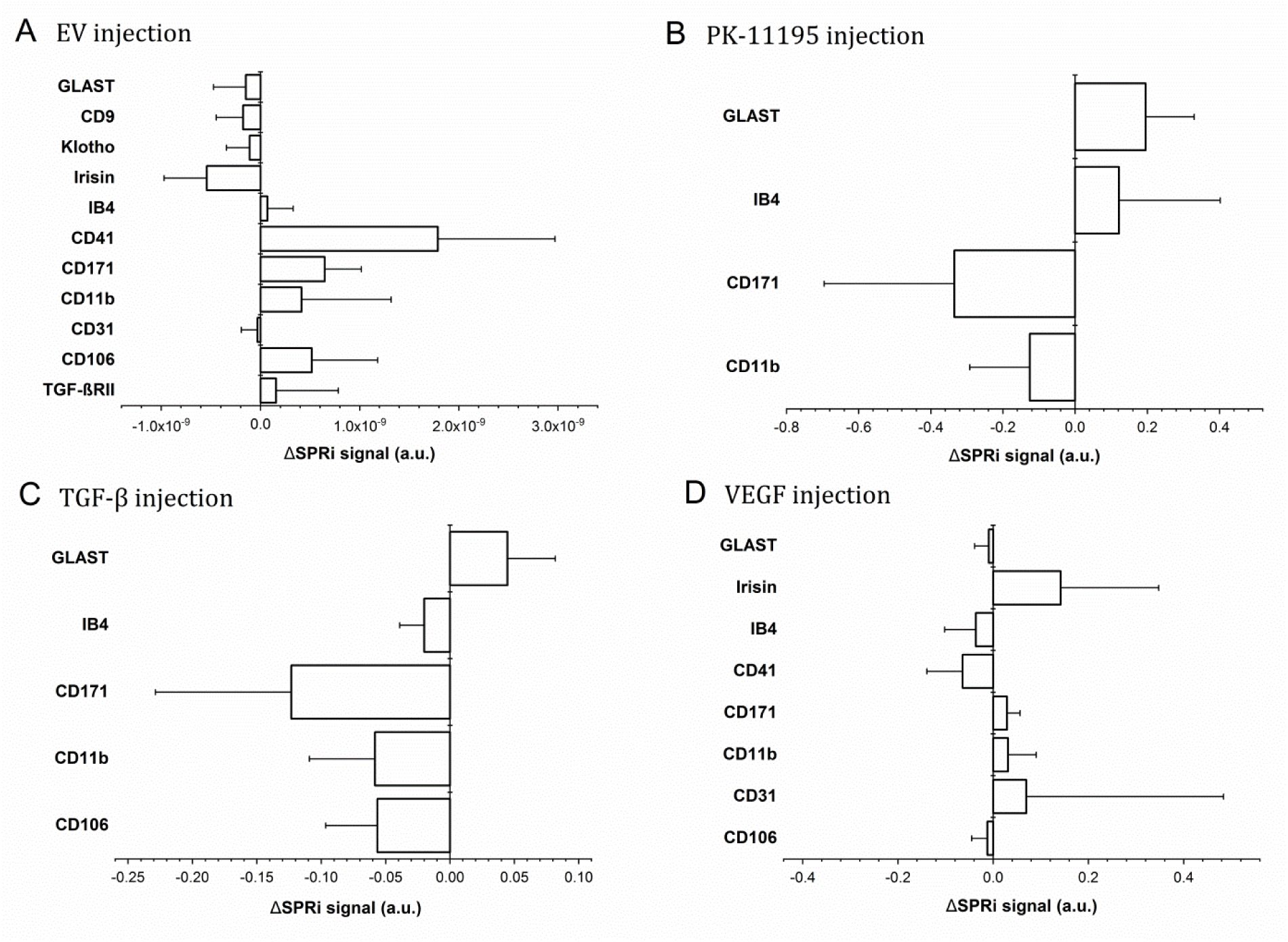
Extracellular Vesicle detection by SPRi biosensor. Bar charts represent the Δ (Δ = T1 SPRi intensity – T0 SPRi intensity) of normalized SPRi signal intensities (average + standard error) collected on the selected ligand spots of EV subfamilies circulating in the serum of people after stroke (*n=*24). Each SPRi experiment consisting of 4 consecutive injections: EV sample (**A**), PK-11195 (**B**), TGF-β (**C**), VEGF (**D**).

To investigate the role of EVs in the neuroinflammatory process after stroke, injection of PK-11195 was performed. PK-11195 is a marker of both acute and chronic cerebral inflammation thanks to its selective binding to the peripheral benzodiazepine receptor (PBR) (also known as the mitochondrial translocator protein or TSPO receptor). An increase of TSPO receptor expression was observed on astrocyte-derived EVs, and a reduction in neuron-derived EVs at T1 (Figure S4A). Regarding microglia-derived EVs, conflicting results were obtained: TSPO was increased on IB4+ (generic microglia) EVs and decreased on CD11b+ (activated microglia) at T1. Nonetheless, for all the considered subfamilies, no statistically significant differences were found (Figure 3B and Figure S4A).

On the same immobilized vesicles, we explored the expression of TGF-βR. After the rehabilitation program, we observed a slight decrement in TGF-βR expression on EVs from microglia, neurons and endothelium, while astrocytes-derived EVs showed an increment of its expression (Figure 3C and Figure S4B).

A final injection of VEGF recombinant protein on immobilized vesicles assessed the involvement of EVs in angiogenesis, neuroplasticity, nerve repair, and glial growth. As expected, the highest SPRi signal intensities were collected on CD31+ EVs both at T0 and T1 (Figure 3D). Considering the Δ of SPRi signals, data showed an increase of VEGF receptor expression on Irisin+ and CD171+ EVs, and a reduction on GLAST+, and CD41+ EVs at T1. Conflicting results were observed on EVs populations coming from microglial and endothelium, with opposite trend between CD11b+ and IB4+ EVs, and CD31+ and CD106+ EVs, respectively. However, for all the considered subfamilies, no statistically significant differences were found (Figure 3D and Figure S4C).

### Correlation study

A partial correlation study was performed (controlling for age, sex, onset-to-admission time) to evaluate the ability of the considered soluble factors and the proposed EV-associated biomarkers to profile ST, describe the severity of the brain lesion, and predict their recovery after rehabilitation, All data are reported in Supplementary Tables.

First of all, we assessed the ability of inflammation biomarkers to describe the clinical status of the patient and to be predictive over the rehabilitation recovery. A positive correlation was observed between the serum levels of IL-6 (Partial Corr: 0.53, *p=* 0.006), TNF-α (Partial Corr: 0.47*, p=* 0.019), and FasL (Partial Corr: 0.55, *p=* 0.005) at T0 and the CIRS score, supporting the role of these cytokines in the description of the general clinical conditions of patients. Moreover, **IL-6 at T0 was shown to negatively correlate with the primary outcome** of our study, i.e. the MBI at T1 (Partial Corr: -0.45, *p=* 0.027), showing the potential of IL-6 in the prediction of functional recovery after rehabilitation.

Interestingly, high circulating levels of NFL at T0 were positively correlated with high pain perceived by patients (NRS) both at T0 (Partial Corr: 0.48, *p=* 0.016) and T1 (Partial Corr: 0.60, *p<*0.001), and with high severity of stroke (NIHSS) at T0 (Partial Corr: 0.47, *p=* 0.015) and at T1 (Partial Corr: 0.56, *p=* 0.002). In addition, NFL at T0 negatively correlate with MBI at T1 (Partial Corr: -0.48, *p=* 0.012), MMSE at T1 (Partial Corr: -0.43, *p=* 0.03), and SPPB at T1 (Partial Corr: -0.48, *p=* 0.027), suggesting that high NFL levels do not only describe the severity of the lesion, but they might be considered predictive of the functional independence of the patients in activities of daily living (MBI) and worse cognitive and motor performances (SPPB), months after the event. Similarly, also the NFH levels at T0 positively correlate with NIHSS both at T0 (Partial Corr: 0.42, *p=* 0.035) and T1 (Partial Corr: 0.44, *p=* 0.026), but not with the other clinical parameters, being a possible marker of subacute lesion severity but not informative about functional recovery (Table S1).

Regarding SPRi experiments, several correlations were found that support the role of EVs in the description of the patient’s clinical status and in the restorative events that are induced by the rehabilitation treatment.

#### Patients’ profiling at admission (T0)

Looking at the correlations between SPRi data and clinical assessment, the role of microglia-and muscle-derived EVs seems to be crucial in the profiling of patients. High levels of CD11b+ EVs are related to better clinical conditions (CIRS) of patients both at T0 (Partial Corr: -0.59, *p=* 0.005) and T1 (Partial Corr: -0.68, *p*<0.001), as well as high levels of circulating neuron-derived EVs (CD171+) that negatively correlate with the global disability (MRS, Partial Corr: -0.52, *p=* 0.048). It is worth mentioning, that multiple correlations were obtained for Irisin+ VEGFR+ EVs (EVs released by muscles cells and responsive to VEGF) and clinical scales of disability and physical performance. Particularly, Irisin+ VEGFR+ EVs at T0 positively correlated with MBI (Partial Corr:0.54, *p=* 0.012), SPPB (Partial Corr: 0.70, *p=* 0.003), and FAC (Partial Corr: 0.74, *p*<0.001) at T0 and negatively correlated with MRS at T0 (Partial Corr: -0.57, *p=* 0.027), suggesting a role of this combined biomarker in the profiling of the disability level and mobility performance of people at rehabilitation hospital admission. Moreover, the circulating levels of TGF-β+ EVs at T0 were correlated negatively with SPPB at T0 (Partial Corr: -0.76, *p*<0.001), indicating that high levels of this TGF-β in EVs at T0 reflects poor motor ability at T0. Finally, the level of astrocyte-derived EVs expressing VEGFR (GLAST+VEGFR+) was found to be the correlated to the cognitive functions of patients (MMSE) at T0, with higher levels of GLAST+VEGFR+ EVs correlated with better performance in the MMSE test (Partial Corr: 0.57, p = 0.014).

In addition, it is not surprising that EVs released by activated platelets sensitive to VEGF (CD41+ VEGFR+) were found to be correlated to the type of stroke event (haemorrhagic vs ischemic; Partial Corr: 0.80, *p =* 0.002) and to the severity of the lesion (NIHSS; Partial Corr: 0.58, *p* = 0.048) (Table S2).

Finally, a negative correlation was observed between NFL (rho= -0.517, p=0.003), IB4+TGF-β+EVs (rho = -0.429, p=0.036) and the MBI at T0 (i.e., **higher levels of NFL and IB4+TGF-β+EVs are associated with a lower MBI score, indicating greater stroke severity and brain damage**). On the other hand, a positive correlation was found between the MBI at T0 and circulating levels of neuronal CD171+ EVs (rho = 0.414, p=0.04), suggesting that **higher levels of circulating neuron-derived EVs could indicate a better health status in ST**. Subsequently, the study of the correlation with the MBI at T1, about two months after the stroke event, showed similar results, with a persistent negative correlation with NFL (rho= -0.498, p = 0.006), but also a new negative correlation with the pro-inflammatory interleukin IL-6 (rho=-0.441, p = 0.021). These results suggest that different biomarkers may influence patients’ functional recovery in different ways over time (Table 4).

**Table 4:** A summary of the correlation study results obtained using machine learning models.

| **Independent Variables** | **MBI at Admission** |  | **MBI at Discharge** |  |
| --- | --- | --- | --- | --- |
|  | *rho* | *p-value* | *rho* | *p-value* |
| Age | 0.044 | 0.817 | -0.206 | 0.283 |
| Sex | -0.224 | 0.235 | -0.155 | 0.421 |
| Stroke Type | **-0.403** | **0.027** | -0.055 | 0.775 |
| **Time enrollment** | -0.267 | 0.154 | **-0.418** | **0.024** |
| **Time rehabilitation** | **-0.825** | **<0.001** | **-0.520** | **0.004** |
| **CIRS** | **-0.362** | **0.049** | **-0.568** | **0.001** |
| **NIHSS** | **-0.756** | **<0.001** | **-0.539** | **0.003** |
| **SPPB** | **0.694** | **<0.001** | **0.681** | **<0.001** |
| **TCT** | **0.861** | **<0.001** | **0.662** | **<0.001** |
| **FAC** | **0.837** | **<0.001** | **0.629** | **0.001** |
| **MMSE** | **0.440** | **0.028** | **0.602** | **0.001** |
| **MRS** | **-0.726** | **<0.001** | **-0.633** | **0.002** |
| NRS | -0.127 | 0.521 | -0.342 | 0.080 |
| HADS/anxiety | -0.468 | 0.079 | -0.509 | 0.053 |
| HADS/depression | -0.236 | 0.398 | -0.416 | 0.123 |
| IL-10 | -0.283 | 0.161 | -0.333 | 0.104 |
| **IL-6** | -0.164 | 0.405 | **-0.441** | **0.022** |
| TNF-α | -0.311 | 0.107 | -0.274 | 0.167 |
| BDNF | 0.247 | 0.206 | 0.134 | 0.506 |
| FasL | -0.228 | 0.252 | -0.195 | 0.341 |
| Leptin | 0.176 | 0.370 | 0.044 | 0.829 |
| VEGFR2 | -0.353 | 0.066 | -0.341 | 0.082 |
| **NFL** | **-0.517** | **0.003** | **-0.498** | **0.006** |
| NFH | **-0.371** | **0.047** | -0.273 | 0.161 |
| CD106+ | -0.051 | 0.808 | -0.228 | 0.274 |
| CD31+ | 0.180 | 0.388 | -0.034 | 0.874 |
| CD11b+ | 0.321 | 0.118 | 0.283 | 0.170 |
| CD171+ | **0.414** | **0.040** | 0.033 | 0.877 |
| CD41+ | -0.121 | 0.656 | 0.372 | 0.156 |
| IB4+ | 0.044 | 0.833 | 0.068 | 0.747 |
| Irisin+ | -0.037 | 0.862 | -0.050 | 0.813 |
| TSPO on CD106+ | -0.022 | 0.918 | -0.048 | 0.818 |
| TSPO on CD31+ | -0.035 | 0.867 | 0.039 | 0.852 |
| TSPO on CD171+ | -0.072 | 0.732 | -0.117 | 0.577 |
| TGF-β on IB4+ | **-0.429** | **0.036** | 0.077 | 0.721 |
| VEGFR on CD11b+ | -0.185 | 0.376 | 0.273 | 0.186 |
| VEGFR on IB4+ | -0.205 | 0.327 | 0.083 | 0.695 |
| VEGFR on Irisin+ | 0.203 | 0.331 | 0.264 | 0.203 |

#### Patients’ profiling at discharge (T1)

As what concerns the profiling of patients at discharge, i.e. after the rehabilitation therapy (T1), high serum concentration of endothelial CD106+ EVs expressing VEGFR was related to improvement of the functional ability of patients (MBI; Partial Corr: 0.45, *p=* 0.04). Moreover, the circulating CD11b+ EVs were confirmed to be correlated with the CIRS scale also at discharge, in line with the above cited observation at admission, whereas the levels of IB4+ EVs (non-activated microglia) at T1 were correlated with higher motor performance (SPPB; Partial Corr: 0.55, *p=* 0.026) and with a reduction of stroke severity (MRS; Partial Corr: -0.68, *p=* 0.004). Finally, the levels of TGF-βR on neuronal CD171+ EVs was negatively correlated with the MBI score (Partial Corr: -0.54 *p=* 0.016), indicating the higher expression of TGF-βR on brain derived EVs describes a considerable disability status (Table S3).

The univariate Spearman analysis also revealed significant associations between clinical scales and the MBI score at discharge. Specifically, better functional outcomes were positively correlated with higher scores on the SPPB (rho = 0.681, p<0.001), TCT (rho = 0.662, p<0.001), FAC (rho = 0.629, p = 0.001), and MMSE (rho = 0.602, p = 0.001), indicating that **preserved motor and cognitive functions are strongly linked to greater rehabilitation gains.** Conversely, negative correlations were observed with CIRS (rho = -0.568, p = 0.001), MRS (rho = -0.633, p = 0.002), and NIHSS (rho = -0.539, p = 0.003), suggesting that **higher comorbidity burden and greater neurological impairment at baseline predict poorer functional recovery**. Additionally, longer time to rehabilitation onset was associated with lower discharge scores (rho = -0.520, p = 0.004), further supporting the **importance of early intervention.**

#### Predictive role of EV biomarkers

Focussing on the predictive potential of EV-associated molecules, the circulating levels of TGF-β+ EVs at T0 were positively correlated with MRS at T1 (Partial Corr: 0.52, *p=* 0.039), indicating that high levels of TGF-β in EVs at T0 (already mentioned as index of poor physical performance) can predict a high residual degree of disability at T1, thus a poor prognosis. In agreement with this observation, higher expression of **TGF-βR on the microglia-derived EVs (IB4+) was found to be predictive of a poor prognosis**, in terms of disability rate and worse functional ability (MBI; Partial Corr: -0.55, *p =* 0.013).

On the contrary, and in line with the previous observation about the role of microglia-derived IB4+ EVs in describing the global disability of patients after rehabilitation, the higher VEGFR expression on IB4+ EVs was correlated to a better physical performance (SPPB) at T1 (Partial Corr: 0.54, *p=* 0.031).

Interestingly, looking at activated microglia (CD11b+ EVs), the expression of the pro-inflammatory TSPO receptor at T0 (CD11b+ TSPO+ EVs) was positively correlated with increased symptoms of anxiety (Partial Corr: 0.91, *p=* 0.011) and depression (Partial Corr: 0.83, *p=* 0.039) at T1.

Considering the astrocyte-derived GLAST+ EVs expressing the surface marker VEGFR (GLAST+ VEGFR+ EVs), they were found to be predictive of better cognitive performance (MMSE; Partial Corr: 0.53, *p=* 0.019), and functional recovery, i.e. lower disability level (MBI; Partial corr: 0.53, *p=* 0.014 and MRS; Partial corr: -0.50*, p=* 0.048), after the rehabilitation program (Table S4).

### ML-based prediction of discharge functional independence

Based on correlation analyses (Table 3) and on previous observations obtained on ischemic stroke patients ^16^, a predictive analysis was conducted using machine learning models.

Biomarkers were included in the multivariate analysis jointly with clinical variables selected based on their significant univariate correlations with the MBI at discharge. SPPB, rehabilitation time, TCT, FAC, and mRS were excluded following a collinearity analysis, as they showed high correlation (r > 0.8) with other retained clinical predictors. The selection was guided by the preference for variables with a lower proportion of missing data, in order to preserve model quality and stability.

The final list of included variables is reported in Table S5 while optimized hyperparameters are reported in Table S6. The variables were selected based on preliminary correlation analysis, previous pilot study ^16^ and expected role in the events that follow the stroke event The SHAP analysis (Figure 4) confirmed the relevance of several T0 variables in predicting discharge outcomes. Among the clinical parameters, the MBI, MMSE score, time from stroke onset to rehabilitation, and NIHSS score emerged as important contributors. Age and comorbidity burden (CIRS) also played a role. Among the biomarkers, key contributors included levels of the pro-inflammatory IL-6, the axonal damage marker NFL, and biomarkers associated with circulating EVs released by activated microglial cells (CD11b+). **High values of IL-6 and NFL, indicators of inflammation and damage, were associated with a lower predicted outcome. In contrast, elevated levels of CD11b+ EVs, along with the overexpression of VEGFR on microglial EVs (IB4+VEGF+ EVs and CD11b+VEGF+ EVs), were associated with a better predicted recovery**

**Figure 4:**
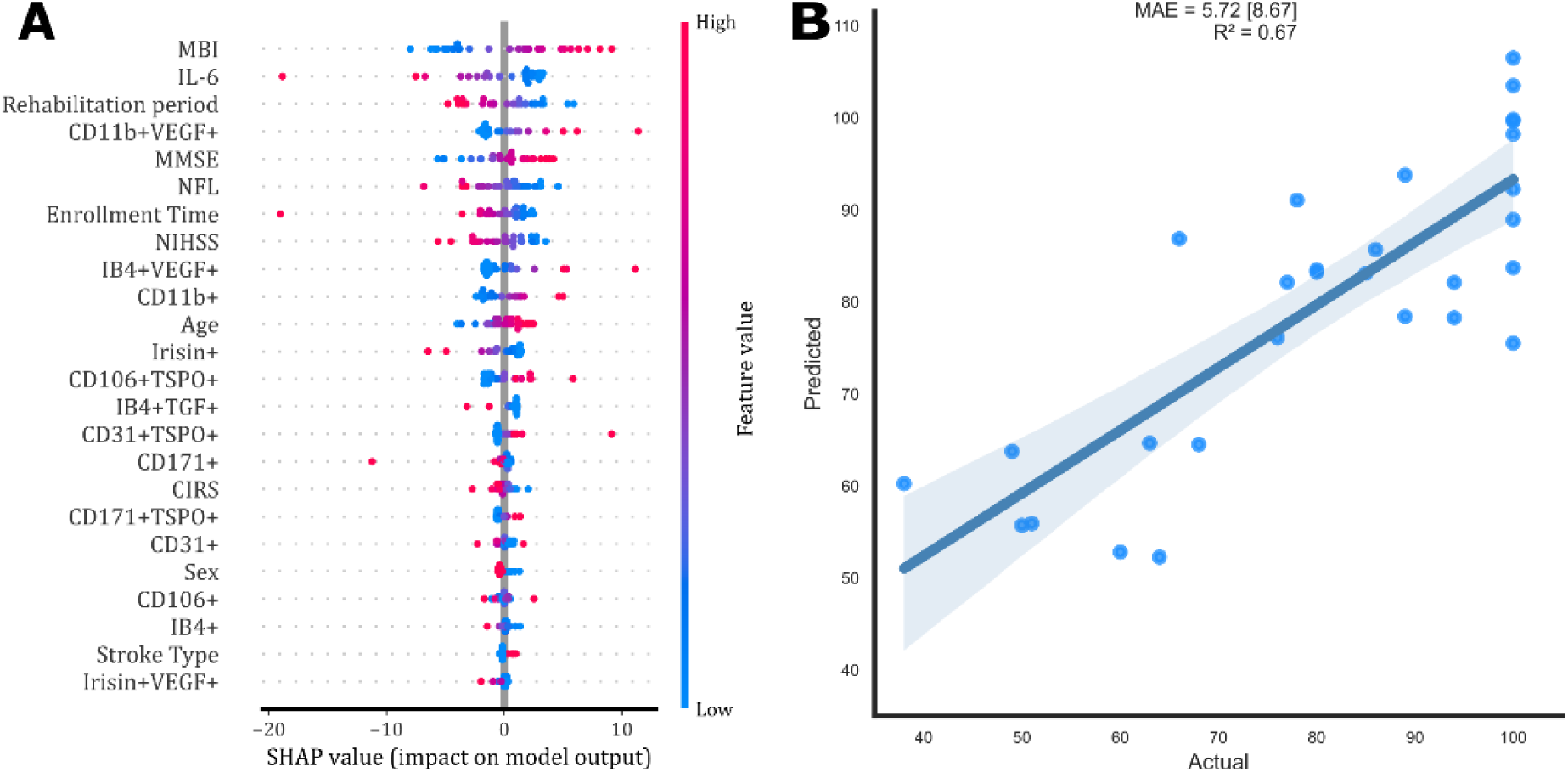
SHAP summary plot of feature importance in the SVR model predicting MBI at discharge (A). Actual versus predicted plot with median absolute error [IQR] and R² coefficient (B).

The accuracy of the obtained cross-validation is 5.72 [IQR = 8.67] points for the estimation of the MBI at discharge, with an R² of 0.67 (Figure 4).

## Discussion

Rehabilitation is the essential therapeutic approach in the management of post-stroke patients, both during the sub-acute and the chronic phase, with the aim to restore totally or partially the cognitive and motor functions lost due to the stroke event. Despite recent innovative advances in rehabilitation medicine, there is still an urgent need for measurable biomarkers to guide personalized rehabilitation intervetions. In the last decades, many inflammatory factors have been explored as prognostic tools, but none have yet entered clinical practice. Some issues are related to i) variability in their temporal profiles of inflammatory markers that decrease after stroke acute phase; ii) patients heterogeneity, iii) lack of standardized the measurement of inflammatory biomarker in terms of timing of sample collection and analysis ^17^.

This study is part of an observational clinical trial conducted in two research hospitals of Fondazione Don Carlo Gnocchi (Italy) and based on a standardized minimal clinical assessment of patients before and after rehabilitation, following the guidelines of the Italian Society of Physical and Rehabilitation Medicine (SIMFER) ^18^. The work expands on previous study reported by the Authors in 2023 ^16^, by integrating clinical, biochemical, and extracellular vesicle (EV) data with AI-assisted predictive modelling.

**The major achievement is the identification of a small panel of** soluble and EV-associated molecules **biomarkers that can support i) the patients profiling in the sub-acute phase and ii) the prediction of the functional recovery after rehabilitation**, defined as improved autonomy in everyday activities measured by MBI score.

Among soluble biomarkers, IL-6 and NFL emerged as key indicators of early clinical severity in the sub-acute phase. IL-6 is known to mediate acute inflammation and to contribute to long-term functional impairment ^19,20^; this study confirms its association with disability (MBI) but not with MRS or MMSE scores as previously reported ^17,19,21^. Similar negative predictive values were obtained for the NFL, structural backbone of neuronal connectivity ^22^. In recent years, neurofilaments have been identified as a sensitive but non-specific marker of neuro-axonal injury in blood and rapidly susceptible to ischemic injury ^23^, even though their blood levels can be affected also by synaptic turnover and secondary neurodegeneration in the late phase after stroke ^24^. NFL levels were reported to vary significantly across three distinct temporal epochs of acute (0–7 days), subacute (9–90 days), and chronic (>90 days) stroke with a steep peak in the early subacute period between 14 and 21 days after stroke, and in general there is a progressive increase within 3 months ^25^. In line with this, we obtained the highest NFL and NFH concentration at T0, and a decrease at T1, r remaining above typical healthy ranges. In this work, high NFL concentration 15-30 days after stroke is associated to lower predictive outcome (MBI), higher stroke severity (NIHSS), lower cognitive performance (MMSE), and lower motor performance (SPPB) at 2 months after stroke. BDNF, TNF-α and IL-10 did not show predictive value, consistent with our previous findings ^16^.

Given the dynamic, multi-cellular nature of post-stroke pathophysiology, the study also investigated circulating EVs. Taking advantage of a multiplexed SPRi-based biosensor, we confirmed that EVs from various cellular sources participate in inflammation, neuroplasticity, and injury response during rehabilitation.

Despite the expected high inter-individual variability, in the considered cohort, a key observation is that rehabilitation impacts on the release of EVs from platelets (CD41+), neurons (CD171+), microglia (CD11b+ and IB4+), and endothelial cells (CD106+). Higher circulating levels of EVs from these cells were detectable in serum at discharge in patients that show better recovery, in terms of stroke severity and cognitive performance.

**One of the major findings is that high serum levels of EVs from neurons (CD171+) and microglia (CD11b+), 2-4 weeks after the stroke event, can yield a positive prognostic value, associated with better recovery at 2 months**. These data suggest the potential role of CD171+ and CD11b+ EVs as biomarkers of good responders in rehabilitation medicine. This might be indicative of active reparative processes but it might also reflect blood–brain barrier alterations that facilitate their passage into circulation.

An even stronger prognostic signal came from the expression of VEGF receptors on EVs. **High levels of VEGF receptor on EVs from microglia, astrocytes, endothelium, and muscle EVs correlated with improved functional, motor, and cognitive recovery**. This aligns with the multifaceted role of VEGF in the neurovascular unit, promoting angiogenesis, neurogenesis, mitochondrial biogenesis, and synaptic plasticity ^26^. Such findings point to VEGF-related EV signalling as a potential mechanism underlying favourable rehabilitation trajectories. However, further studies are needed to investigate the functional role of VEGF on brain cells after stroke.

In the considered cohort, **microglia-derived EVs showed a particularly marked prognostic relevance**. Specifically, elevated levels of CD11b+ EVs and the overexpression of receptors for VEGF on microglial EVs (IB4+VEGF+ EVs and CD11b+VEGF+ EVs) were associated with a better outcome, whereas the increased TGF-β binding by IB4+ EVs (TGF-β+IB4+ EVs) was associated to worse clinical status. This supports the well-known dual nature of microglial responses, oscillating between neuroinflammatory and reparative functions ^27^. Moreover, it’s well established that TGF-β, especially isoform 1, is universally induced by acute and chronic brain injury, including stroke ^28^. TGF-β receptors are also present in all major cell types and activated microglia and macrophages are the predominant sources of increased TGF-β after stroke ^28^. Actually, in literature, other studies underline the pleiotropic and context-dependent biological effects of TGF-β signalling in stroke, influenced by both injury type and timing. Given the broad and context-dependent roles of TGF-β in brain injury, elevated TGF-β+ EVs may reflect persistent glial dysfunction in the subacute and chronic phases.

Regarding EVs from skeletal muscle, the expected increase in Irisin+ EVs after motor rehabilitation was not observe although we report an increased ability of Irisin+ EVs to bind VEGF. These data are in line with the controversial literature regarding the increased release of Irisin after exercise by skeletal muscle due to the impact of exercise regimen, mode of exercise, time point of sampling, and study population reported ^29^. Besides, based on previous literature ^16^, CD31+ EVs from the neurovascular unit were expected to have an impact on the rehabilitation outcome, nonetheless in the present study the variations in circulating endothelial EVs was not correlated with patients’ recovery. Despite being tested on a limited patient sample and thus warranting cautious interpretation, the nonlinear predictive models employed to estimate functional recovery reinforced the prognostic value of biomarkers, yielding accuracy levels aligned with previous literature ^30–33^, and potentially useful for informing rehabilitation planning.

The results of the present study represent a milestone in the identification of predictive biomarkers for rehabilitation recovery after stroke. Despite limitations related to the number of recruited subjects and to the considerable inter-individual variability of data, partly due to the heterogeneous nature of the stroke population, our data confirm the crucial role of neuroinflammatory mediators in the restorative events that follow the stroke lesion. The use of a sensitive multiplexed biosensor has allowed us to test the involvement of multiple brain and non-brain cells in the rehabilitation induced mechanisms that follow a brain lesion, supporting the use of EV-associated molecules as predictive biomarkers of stroke recovery. Although future studies are needed to validate our findings in a wider cohort of subjects, our results offer a robust starting point for upcoming research into the long-term mechanisms of stroke recovery and the development of therapeutic strategies to support brain remodelling.

## Conclusions

In conclusion, the major finding of the present study is the identification of a panel of few circulating biomarkers for the profiling of the clinical status of stroke patients in the sub-acute phase of the pathology and for the prediction of their response to rehabilitation treatment. Specifically, microglia-derived EVs - detectable in blood - proved to be valuable tools to objectively assess the neuroinflammatory status of stroke patients and to predict their functional recovery after rehabilitation.

## Data Availability

The datasets generated during and/or analyzed during the current study are available from the corresponding author on reasonable request.

## Ethics approval and consent to participate

The present study was approved by the Ethical Committee of Fondazione Don Carlo Gnocchi Onlus ((protocol number: 20/2018/CE_FdG/FC/SA).), and carried out according to the Declaration of Helsinki.

## Authors’ contributions

A.G., M.B. and S.P. conceived this project and designed the study; Au.M. performed most of SPRi experiments; S.P. and A.G. supervised the experiments; P.A., D.B., L.C., F.C., and J.S.N. were responsible for patients’ recruitment and clinical assessment; P.P. performed the AFM experiments; L.R., E.M. and An.M. performed AI-based analysis; Au.M., S.P., and A.G. performed data analysis; A.G. supervised the study; M.B. and A.G. were responsible for acquisition of the financial support for the project; Au.M. and A.G. wrote the original draft; all authors contributed to manuscript review.

